# Behavioral Risk Identification and Decision Guidance for Engagement (BRIDGE): Research protocol for an evaluation of an HIV treatment retention toolkit for the early treatment period

**DOI:** 10.64898/2026.03.24.26349199

**Authors:** L Sande, M Maskew, N Mutanda, E Kachingwe, A Morgan, V Ntjikelane, S Chiwaye, M Benade, AR Marri, L Malala, M Manganye, S Rosen, N Scott

## Abstract

**Background:** Interruptions in HIV care pose a major challenge to achieving HIV control goals in many countries, with 30% of clients who initiate antiretroviral therapy (ART) in South Africa experiencing an interruption of >28 days during their first six months on treatment. South Africa introduced revised guidelines in 2023 to improve outcomes during this early treatment period, but guideline compliance remains incomplete and gaps in the support provided to both clients and providers to optimize service delivery and health outcomes.

**Protocol:** BRIDGE (Behavioral Risk Identification and Decision Guidance for Engagement) is a mixed-methods evaluation of a package of light-touch, low-cost interventions aimed at improving both clients’ and providers’ experiences of care, increasing compliance with the 2023 guidelines, supporting clients to remain in care, and ultimately reducing the incidence of missed visits during the early treatment period. Components of the BRIDGE Retention Toolkit include an intervention navigator to help clients self-assess areas of vulnerability for disengagement from care and identify appropriate interventions; client roadmap to explain the treatment journey for the early treatment period; WhatsApp-based counseling tool for clients; guideline reference for providers; and tracing job aids. The tookit will be piloted at 6-8 public sector primary health facilities for a one-month period. The primary outcome will be the probability of returning less than 28 days late for the next scheduled clinic visit, assessed using electronic medical record data for the pilot and comparison sites. Pilot outcomes will be compared to both their own probabilities prior to the pilot and to probabilities from comparable non-pilot facilities. Implementation outcomes to be assessed using qualitative interview data from both clients and providers will include reach, implementation fidelity, adoption (uptake), costs, feasibility, appropriateness, and acceptability.

**Discussion:** The evaluation will assess the implementation and preliminary effectiveness of a set of interventions designed to improve client outcomes during the early HIV treatment period. If some or all of the BRIDGE tools are found to be helpful and/or are associated with a reduction in missed clinic visits, they will comprise a readily scalable and affordable intervention to help address a major barrier in large-scale HIV treatment programs.

## Introduction

In South Africa, an estimated 78% of the roughly 8 million people living with HIV are on antiretroviral therapy (ART) [1]. The main challenge to increasing this proportion is keeping people in care after treatment initiation. The problem of disengagement from HIV care within the public sector in South Africa has been well documented [2–4]. This problem is at its worst during the early treatment period (first six months after ART initiation or re-initiation), with only an estimated 59% of those starting treatment remaining continuously in care, with no interruptions greater than 28 days, by the six-month point[4]. The probability of missing one’s next scheduled clinic visit during this period approaches 9% for the visit after initiation; 30% of clients experience an interruption during the first six months[4,5].

Major improvements to HIV treatment delivery programs that have been adopted in South Africa and other countries in the past decade, including universal ART eligibility, same-day ART initiation, and differentiated service delivery models, have generally passed over the early treatment period, which for most clients continues to require frequent clinic visits and a one-size-fits-all model of care that is not tailored to clients’ needs or preferences [6]. Only recently has attention focused on the high rate of interruption and disengagement among those starting treatment, who have no yet achieved viral suppression or settled into long-term patterns of care[6–8].

The probability of disengagement from HIV care during the early treatment period is influenced by characteristics of healthcare facilities, healthcare providers, treatment clients, and communities[9–11]. At the facility level, this includes quality of care provided, availability and use of facility-based retention interventions, clinic volumes and staffing resources available, and location and accessibility of the facility site. At the health provider level, perceptions of patient risk of disengagement, time available for each consultation, working conditions, interpersonal skills, and awareness and knowledge of guideline recommendations and retention interventions could also impact the patient-provider encounter. At the client level, perceived lack of benefit of ART, negative side effects of medication, stigma, lack of social support, negative experiences with health facilities and staff, financial constraints, and work commitments may make adherence to visit schedules challenging. Proximal life events such as unexpected mobility, health issues, forgetting an appointment date, or other social responsibilities can also contribute to missed visits[12]. Finally, stigma remains a major contributor to disengagement from care at all levels[10].

In 2023, the South African National Department of Health (NDOH) revised its HIV care and treatment guidelines and treatment adherence guidelines to better address some of the concerns and barriers mentioned above. Key changes pertaining to the early treatment period included moving the first scheduled viral load test from six months after initiation to three months; allowing intervals of 6 months between clinic visits and enrollment in differentiated models of care (DMOCs) at the four-month point for clients whose three-month viral load was suppressed; and reducing the recommended number of clinic visits in the first year of treatment from 11 to 6. These revisions, which aimed to make treatment less burdensome for both clients and providers, constituted significant changes in the clinical management of clients starting or re-starting ART and required providers to incorporate different schedules and procedures into their routine consultations. Although introduced in April 2023, facility-level compliance with the guidelines is believed to remain incomplete.

The Retain6 study (www.sites.bu.edu/ambit) was designed to generate evidence about when and why clients disengage from or interrupt treatment during the early period, understand the barriers to continuous engagement in care during this period, and identify feasible opportunities to improve outcomes. In South Africa, findings from the first four years of Retain6[10,13–15] have led to the development of a package of light-touch, low-cost interventions aimed at improving both clients’ and providers’ experiences of care, increasing compliance with the 2023 guidelines, supporting clients to remain in care, and ultimately reducing the incidence of missed visits during the early treatment period. Here we describe the Behavioral Risk Identification and Decision Guidance for Engagement (BRIDGE) study’s intervention package and evaluation plan.

## Protocol

BRIDGE is a mixed-methods evaluation of a “retention toolkit” designed to improve experiences and outcomes during the first six months of ART. Most of the tools in the toolkit are being designed for BRIDGE, in collaboration with government and facility stakeholders and ART clients; the toolkit will also adopt one existing digital app. The toolkit will be implemented by facility staff for roughly a one-month pilot period at 4-8 facilities that provide routine HIV care. It will be evaluated through qualitative interviews with clients and staff and quantitative analysis of facility-level client data, with outcomes that include the incidence of missed visits as well as the uptake, feasibility, acceptability, and cost of the interventions in the toolkit. The full study protocol is provided as Supplementary file 1.

### Objectives

BRIDGE has three main objectives. First, we aim to develop a retention toolkit using a co-design process in collaboration with key stakeholders, including Department of Health officials, facility healthcare providers, ART clients, and external experts on HIV treatment service delivery. The toolkit is intended to fill specific needs identified by providers and clients and to be implementable using existing human and other resources at typical public sector primary healthcare clinics (PHCs), at little additional cost to the Department of Health. Second, the toolkit will be piloted at selected PHCs for an implementation period of approximately one month. Implementation will be carried out by existing facility staff as part of routinely service delivery during the pilot period. And third, we will conduct a mixed-method evaluation of the retention toolkit based on the pilot experience. The evaluation will include both i) process outcomes such as implementation fidelity, consistency of use, resource requirements, acceptability to providers and clients, client satisfaction with their experience; and ii) the impact of the toolkit on patient visit attendance during and immediately after the pilot period, compared to the period prior to toolkit implementation.

Based on results collected earlier in Retain6, much of BRIDGE focuses on gaps in treatment literacy, lack of understanding of the treatment experience, and social support for clients and on guideline awareness and compliance support for providers, issues that can potentially be addressed through light-touch, low-cost approaches. For practical purposes, we selected tools that we believe can be implemented at very little additional budgetary cost to providers or clients and requiring minimal additional time beyond current needs.

### Toolkit components

The BRIDGE retention toolkit has five components. Each component is being co-developed collaboratively with stakeholders as part of the project, and we anticipate that we will continue to revise the tools over the course of the pilot period as we receive feedback from users. Each tool aims to address a concern raised by providers or clients during surveys, interviews, and focus groups conducted earlier in the Retain6 project.

We describe the current draft of each tool below; a summary is provided in Table 1. (As explained below, revision of the tools will be iterative, throughout the orientation and pilot periods.)

**Table 1.** Summary of the BRIDGE retention toolkit components.

| Component | Format | Use | Staff cadre | Use schedule |
| --- | --- | --- | --- | --- |
| i. Intervention navigator | Handout for clients; poster to be displayed in each consultation room and/or in counselling rooms and waiting areas, as determined by the facility. | Maps self-identified barriers/risk factors directly to potential solutions. Give to each ART client in the waiting area at reception or during consultation. Client can review while waiting for consultation (can write on it (tick off the boxes) if desired). May be reviewed during group information sessions in waiting area. Provider reviews it with client during consultation and suggests intervention/support strategies. Client takes home if desired. | Client, counselor, and nurse | Initial visit; later visits during early treatment period as needed by clients |
| ii. Client roadmap | Handout for clients; poster for wall in each consultation or counselor's room | Give to each initiating or re-initiating client in the counselling or consultation room. Review with providers and fill in next visit date(s). Client takes home if desired. | Client and nurse or counselor | Initial visit; later visits during early treatment period as needed by clients |
| iii. WhatsApp Health Coach | Posted in waiting areas and consultation rooms | Call clients' attention to the QR code that they can use to access the app. Refer clients to poster during clinical consultation. | Nurse or counselor | As accessed by clients |
| iv. Guideline reference poster | Posted in each consultation room and laminated desk reference taped to desk | Use before, during, and after client consultations to confirm all steps have been completed. | Nurse | As needed by providers |
| v. Tracing support tools | Handout for all tracing staff | Use before starting to trace any client to confirm all steps have been completed. | Counselor/ WBOT/ CHW | Every tracing action |
|  | Stickers to keep in pile at reception desk. | Update client contact details at every visit. | Clerk or nurse | Every client visit |

WBOT, ward-based outreach team; CHW, community health worker

#### i. Intervention navigator

One of the major challenges encountered during the early treatment, from ART initiation onward, is how best to target available support and interventions to clients who need them, without adding to the care burden of or investing scarce resources in those who do not need support. We initially sought a strategy to allow providers to score individual clients’ risk of missing their next clinic visit, with the intent to create a “risk triaging algorithm” that providers could then utilize to classify individual risk levels and thus determine what types of support should be offered[16]. Consultation with stakeholders, including providers and program managers, however, led us to focus on asking clients to determine their own potential vulnerabilities and consider options for addressing these vulnerabilities, while offering providers assistance in matching specific vulnerabilities to appropriate interventions.

For BRIDGE, we developed a combined self-assessment/intervention mapping tool called the Intervention Navigator that can be used by both clients and providers to guide identification and discussion of potential risks or vulnerabilities and the interventions or support that a client might choose. The tool (Figure 1) groups factors that could lead to disengagement from care into four categories: treatment readiness and emotional/psychological fears; obstacles in attending clinic visits; other health concerns that may complicate HIV care; and difficulty remembering treatment or visit schedules. Each category is linked to one set of potential steps clients themselves can take to address these barriers and to one set of services potentially available from the clinic (the range of interventions available varies somewhat by facility).

**Figure 1.**
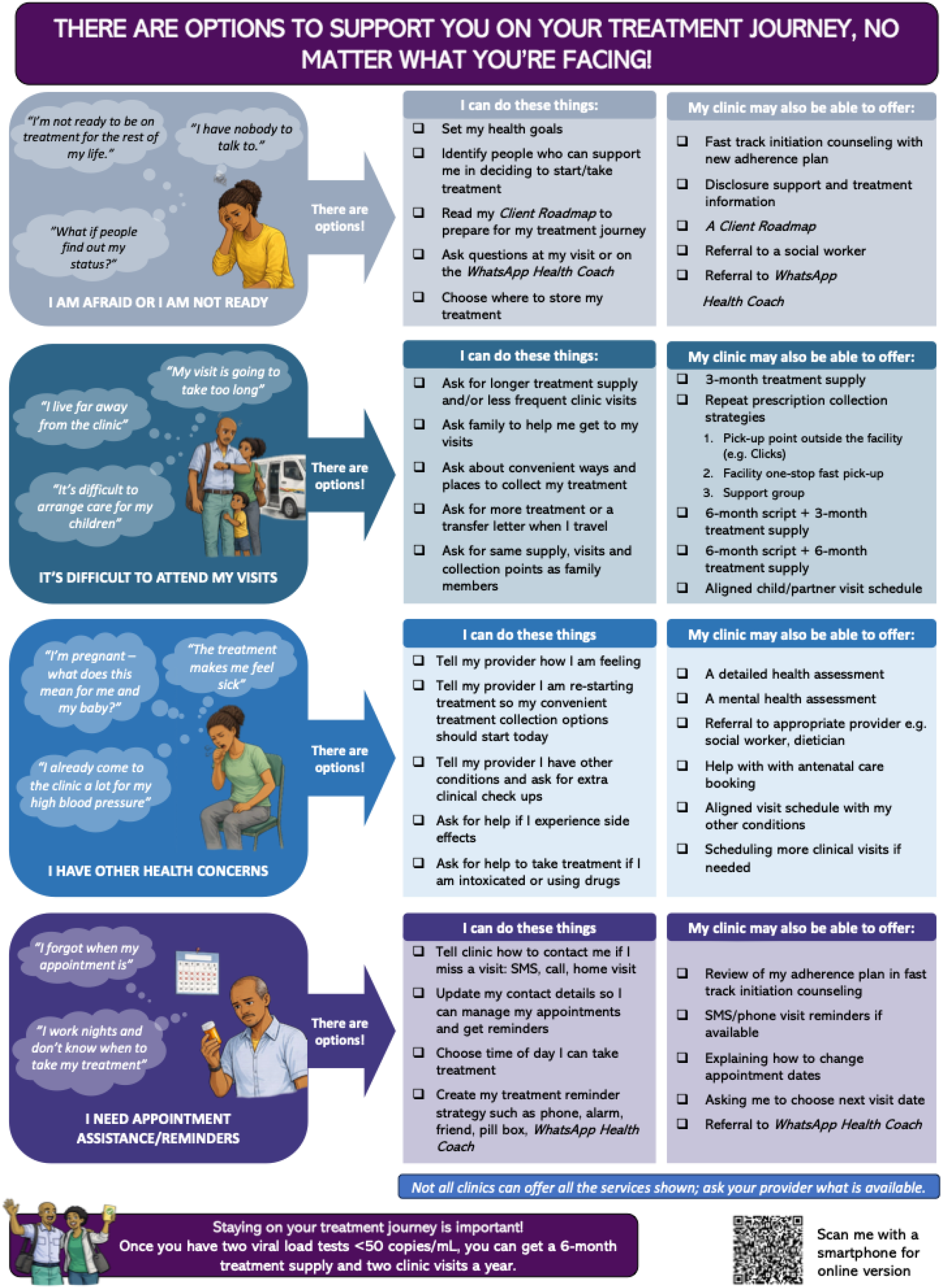
Intervention navigator.

The tool will be handed out to individual clients in the waiting area and/or during clinical consultations or counseling appointments to guide conversation and solutions to overcoming barriers to care. It will also be provided as a laminated poster that can be hung each consultation and counseling room, for easy reference during consultations and group information sessions.

#### ii. Client roadmap

One of the findings of Retain6’s surveys and interviews was that clients starting ART do not fully understand the pathway they will follow once they are on treatment. They fear having to visit the clinic monthly for the rest of their lives, and the potential to enroll in a low-intensity model of care, receive multiple months of medication at once, and/or obtain medications from a more convenient pickup point is not clear. Other guidance, such as what to do if a visit is missed or what to do in case of a problem, may also be not be understood or retained. To address this barrier to returning for one’s next visit(s), we developed a “roadmap” handout to distribute to clients that provides a visual aid for counselors and clinicians as they explain the treatment process. Such roadmaps are a common tool for supporting patients with chronic diseases[17], but they are not currently in use for HIV care in South Africa.

Development of the client journey roadmap began with an illustration of the steps outlined in the treatment guidelines. We then elicited input from a wide range of stakeholders, including health systems experts, health policy makers, providers, and patients, about the content and design of the roadmap, ending up with a tri-fold brochure with the map illustration on one side and user-friendly information for the client on the other side (Figure 2). The roadmap provides spaces for filling in upcoming visit dates and for notes from the patient or provider. A paper copy of the roadmap will be given to all clients who are initiating or in their first six months of treatment, and a poster of it will be displayed in consultation and counselling rooms. We anticipate that future versions may include one or more QR codes to allow users to link to relevant websites as well.

**Figure 2.**
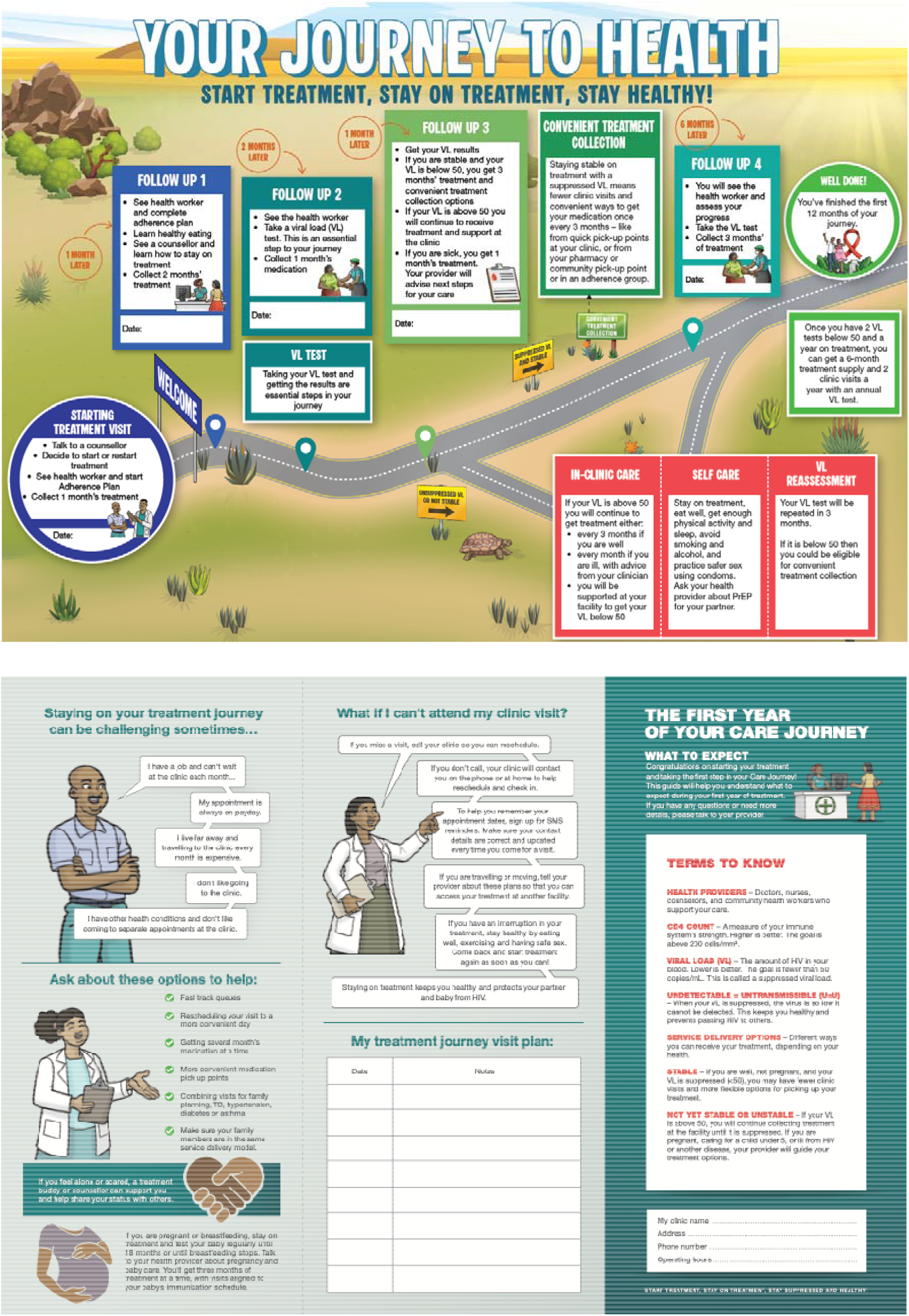
Client roadmap.

#### iii. WhatsApp® Health Coach

Many clients participating in the Retain6 surveys and focus groups indicated that they would like more counseling and treatment literacy support than they received during the early treatment period. At a time when staff resources are scarce and most efforts are directed at making service delivery less intensive, rather than more, offering additional in-person counseling or education is not feasible. Instead, the BRIDGE toolkit will refer clients to an existing app-based service developed by another organization working in South Africa, Population Services International (PSI). The app, called the WhatsApp Health Coach, is an AI-powered digital health tool designed to provide tailored, on-demand health information, advice, and support to people living with or vulnerable to HIV and TB, complementing existing facility-based care. Inspired by South Africa’s successful Coach Mpilo peer-support model[18] the AI Coach replicates key functions of real-life health coaches—offering confidential, user-driven, text-based guidance through WhatsApp. Users can engage the chatbot at any time to ask questions about HIV, TB, sexual health, and related psychosocial issues, receive reminders for medication or clinic visits, and access information on nearby healthcare services.

Users access the AI Coach through a QR code or WhatsApp prompt and interact voluntarily with the coach, including the option to request linkage to care or escalate to a real-life nurse coach for complex or high-risk issues. It can be accessed anonymously by those who prefer not to provide identifying details. BRIDGE will give clients at our pilot clinics access to a QR code that will take them to the WhatsApp Health Coach, where they can follow instructions to enroll and then access the app at any time during the early treatment period, whether or not they remain formally in care or in contact with the facility, offering a resource for ongoing, self-managed support (Figure 3).

**Figure 3.**
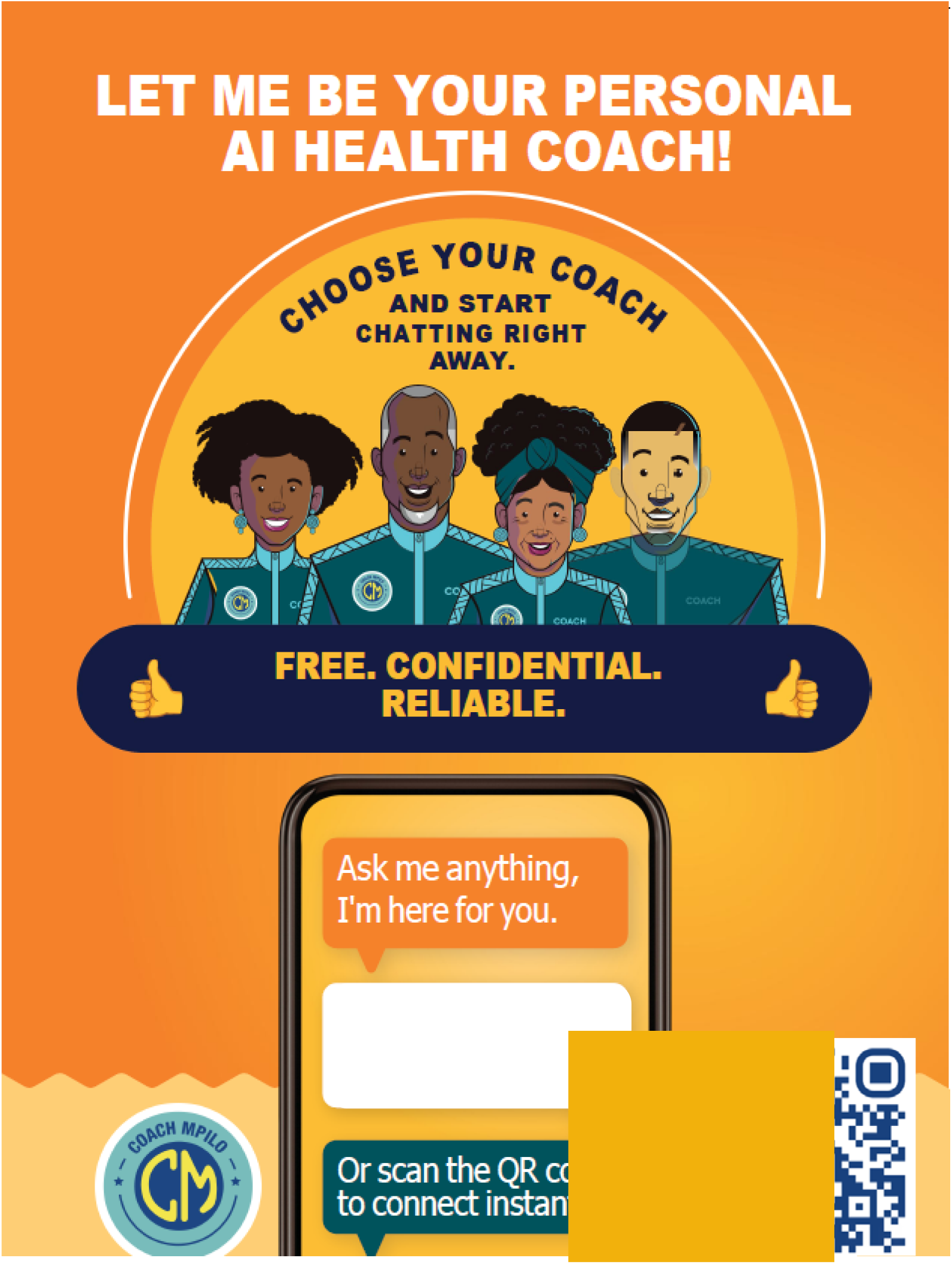
WhatsApp Health Coach.

The WhatsApp Health Coach has no cost to users but does require internet or mobile data access or cellular airtime for utilizing WhatsApp. More information about this tool is available from https://cquin.icap.columbia.edu/wp-content/uploads/2025/11/Malone_14c_Final.pdf.

#### iv. Guideline reference poster

Retain6’s analysis of compliance with the provisions of the 2023 ART guidelines that pertain to the uptake and timing of viral load testing and enrollment in DMOCs and 6-monthly clinic visits suggest very incomplete implementation, with substantial variance in adoption by facility. In Gauteng Province, for example, in a sample of 1,416 clients whose viral load was suppressed at their 3-month test, only 11% were documented as enrolled in a DMOC at four months. Another 40% enrolled at or after 6 months, reflecting continued reliance on previous treatment guidelines.

Although all providers have some access to both hard and online copies of the guidelines, the hard copy documents may not be available in consultation rooms and providers may not have sufficient time or internet access to consult the guidelines online while interacting with clients. Familiarity with the lengthy guideline documents may also be limited. To assist providers to follow guideline schedules and procedures, BRIDGE includes a reference poster that summarizes the key elements of the guidelines and highlights specific pages of the main guidelines document so that a provider can more readily access the relevant information (Figure 4).

**Figure 4.**
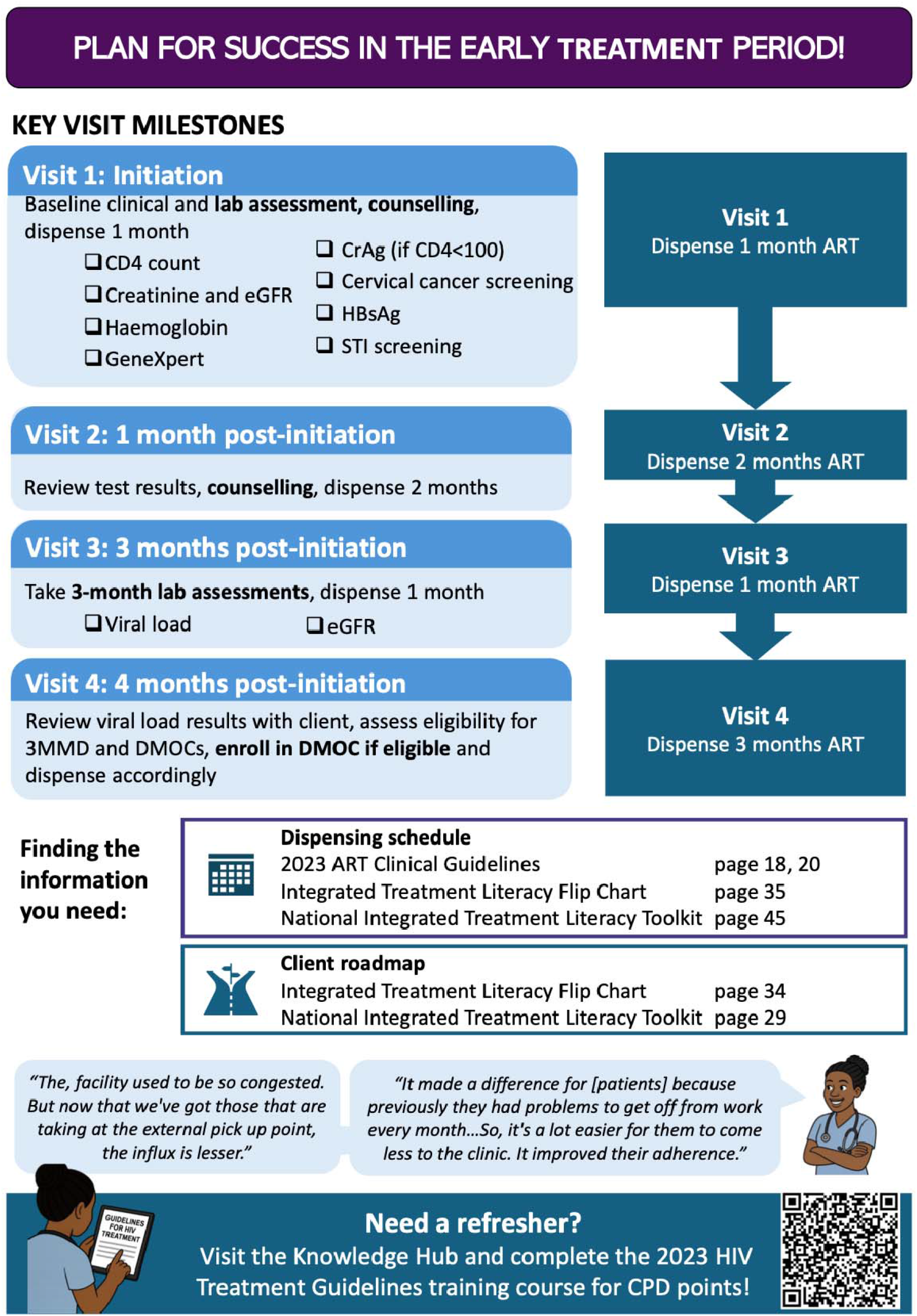
Guideline reference poster.

#### v. Tracing support tools

As mentioned above, interruptions to care (missing scheduled clinic visits and medication refills) are extremely common during the early treatment period. The most common response of facilities to missed visits is to attempt to contact the client through a process labeled in the guidelines as “tracing and recall.” Guidelines recommend that tracing staff (typically counselors and community health workers) telephone clients who are 7 or more days late for a scheduled visit. If the client cannot be reached by phone, a home visit is recommended. The effectiveness of tracing in leading clients to return to care is uncertain, with widely varying results reported in the literature[19]. Facilities report that a major impediment to the success of tracing is not having current contact details available in clients’ files. Prior research by Retain6 also found that due to incomplete data capture, many clients at our study facilities who should be traced are either not identified or not reached, while many who do not require tracing are contacted[20].

To try to improve the efficiency of tracing clients who interrupt care, while maintaining fidelity to guidelines, BRIDGE will introduce two tools affecting the tracing process. The first is a job aid for staff who conduct tracing. The job aid provides a detailed, step-by-step process for identifying clients to be traced, confirming their eligibility for tracing, conducting the tracing, and recording results (Figure 5a). The second is a pre-printed sticker/adhesive note form that will be attached to client files and serve as a reminder to the receptionist or other intake staff that contact details should be updated and confirmed at every visit, rather than only at the first visit as is currently most common (Figure 5b).

**Figure 5.**
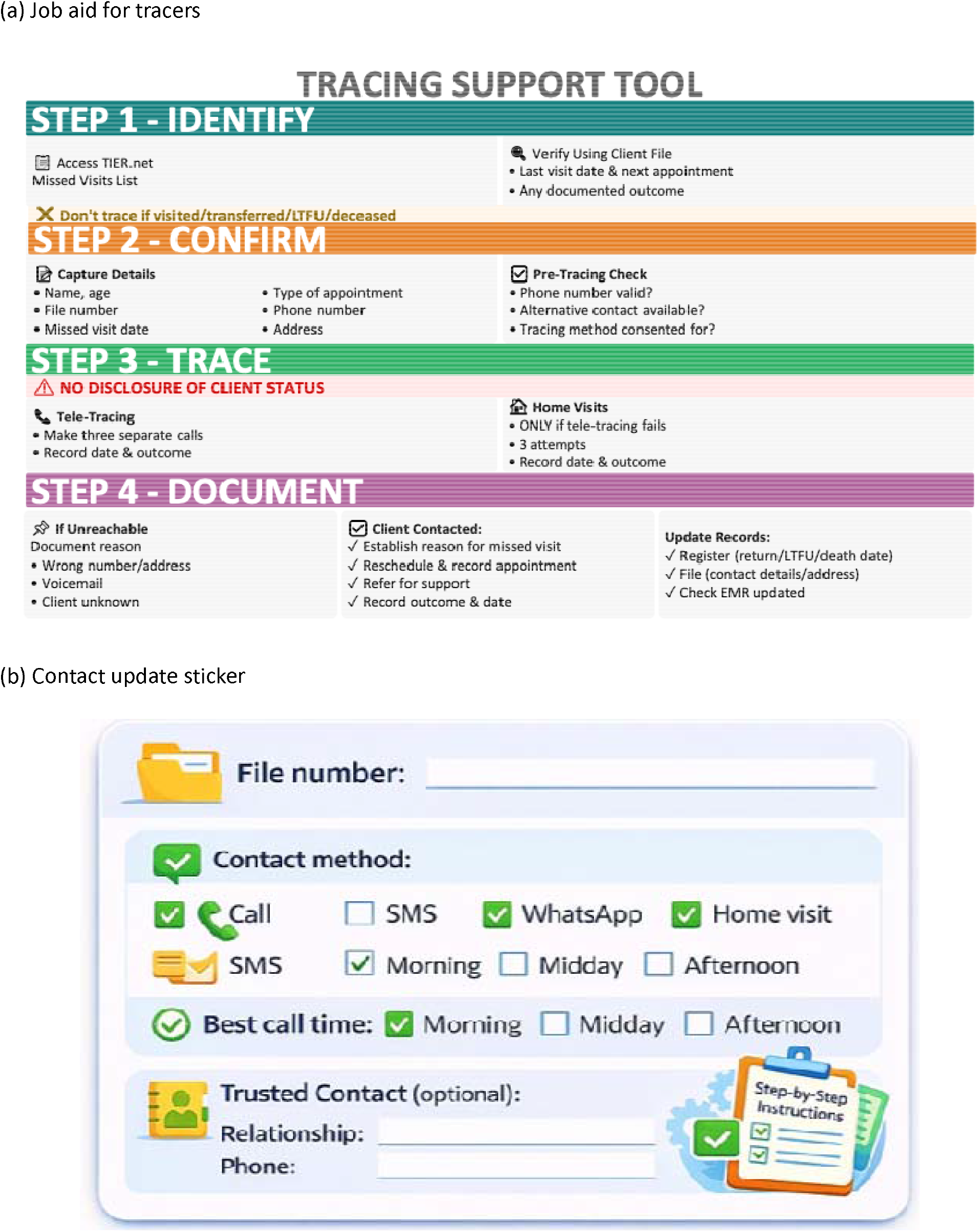
Tracing support tools. (a) Job aid for tracers. (b) Contact update sticker.

### Pilot implementation

The BRIDGE retention toolkit will be piloted at between six and eight primary health clinics in Gauteng Province and in KwaZulu Natal (KZN) and/or Mpumalanga Provinces, depending on the timing of provincial permissions. The sites are among those participating in the SENTINEL network created by the AMBIT project to monitor changes to service delivery, uptake, and outcomes over time[21].

At each site, clinic staff (not study staff) will utilize the tools, as summarized in Table 1. The pilot is expected to be conducted for four weeks at each site prior to evaluation. (While study materials will be donated to the sites for ongoing use when the pilot has been completed, project resources allow only a one-month implementation period by the study itself).

Following ethics review and approval, permission to conduct the pilot at the study clinics was obtained from provincial and district health offices. After explaining the project to the facility, operations, or ART manager at each site, the BRIDGE toolkit will be introduced to relevant healthcare providers (nurses, counselors) during a three-hour orientation session conducted by the research team. The orientation will explain the origins of the toolkit and then review and demonstrate the use of each tool. Because the study sites for the pilot have previously participated in the research activities that led to BRIDGE, staff are familiar with the research team and the concerns we are addressing. During the orientation, input from potential users (staff) at each site will be sought as part of the co-creation process. The orientation session will also consider practical implementation details, such as which staff cadres should be responsible for handouts, who should fill in the updated contact form, and where posters should be hung.

After the orientation, each study site will be asked to utilize the toolkit for all eligible clients for a period of four weeks. Implementation will be carried entirely by facility staff, with the study team providing support but not directly participating. Standard operating procedures will be developed and utilized by the study team at each site, providing a benchmark against which we will assess implementation consistency. A study coordinator and/or research assistant will visit each site 2-3 times per week during the pilot period to ensure that supplies of handouts remain sufficient, restock materials as needed, answer questions, elicit comments and feedback from the staff who are using the toolkit, and provide any other support that may be requested. In addition, each implementing provider will be provided with a “toolkit cheat sheet” to remind providers of how to use each tool. This document will be a user-friendly version of Table 1 above.

For logistical reasons, the start of the one-month pilot period will vary slightly from site to site. Lessons learned during the first two weeks of implementation at the earliest sites to launch the pilot will be incorporated into the tools as implementation progresses, with updated versions distributed for use whenever available.

### Data collection

We will collect three types of data to evaluate the BRIDGE retention toolkit. First, we will use electronic medical record (EMR) data captured by TIER.Net, South Africa’s HIV EMR to estimate the primary outcome of return within 28 days of the next scheduled visit. EMR data will be extracted for all eligible ART clients making visits to the study sites during the six months prior to pilot implementation, for the duration of the pilot, and for up to six months following the pilot. Parallel EMR data will also be extracted for all clients meeting eligibility criteria at up to 9 of the remaining non-pilot facilities in the SENTINEL network in the same provinces as the pilot facilities, for use as comparison sites. Fields to be collected include sex, age, date of ART initiation, dates of all scheduled and observed clinic visits and medication pickups, and dates and results of HIV-associated laboratory tests. A waiver of informed consent has been granted for EMR data use.

Second, we will conduct telephone interviews with clients in their first 12 months of treatment who were exposed to at least one component of the toolkit. Clinic staff will inform clients with whom they used one or more components of the toolkit about the study and refer interested clients to study staff stationed at the facility. In a private location in the facility, study staff will describe the aims and overview of the study. Willing and eligible participants will be asked to provide written informed consent to participate in the study and will then be asked for a preferred telephone number and date and time that they wish to be contacted; interviews will be scheduled for approximately one week after enrollment or soon thereafter. Study staff will then call the clients, confirm they are in a private space, and proceed with a short semi-structured telephone interview. The interview will ask about component(s) of the toolkit that the client was exposed to, acceptability, and suggestions for improvement. The interviews will last no longer than 30 minutes. Interviews will be audio recorded, transcribed, and translated as necessary.

Finally, we will conduct in-person interviews with providers to explore their experiences with and views on the tools after using them for one month. Study staff will approach eligible facility staff who had been working in the ART clinic during the pilot implementation and solicit written informed consent prior to the interviews. We will collect basic descriptive information (e.g. current position, number of years in current position, number of years working in HIV program) and then use a structured interview guide to ask providers about their perspectives on the tools. We will ask about perceived benefits of the tools, things that may make it difficult to use the tools, and suggestions for improvement. Interviews will be audio recorded and study staff will also take written notes. We anticipate that each survey and interview to take no more than 60 minutes to complete.

### Evaluation of the pilot—primary quantitative outcome

BRIDGE will be evaluated for preliminary clinical outcomes and process outcomes. The primary clinical outcome to be assessed is the probability of returning less than 28 days late for the next scheduled clinic visit. This will be measured using routinely collected, electronic medical record (EMR) data captured by TIER.Net, South Africa’s HIV EMR. Clients who attend a visit at a participating facility during the pilot implementation period will be considered exposed to the retention intervention, while those who attended visits before or after pilot implementation or at non-pilot sites will be considered unexposed. We will compare the aggregate proportion of ART clients in their first six months on treatment who are ≤28 days late for their next clinic visit at implementing facilities to proportions of visit attendance at non-implementing facilities during the pilot implementation period. We will also compare visit attendance rates among clients at implementing facilities during the implementing period to visit attendance rates at the same facilities in the six months prior to implementation of the toolkit.

For the primary clinical outcome, the analytic approach will start with a description of the demographic and clinical characteristics of clients making ART-related visits at participating sites stratified by exposure group. Next, we will describe the baseline prevalence of late (>28 days late for a scheduled date) visit attendance stratified by exposure group. We will summarize these with simple proportions and estimate the effect of exposure to the toolkit on visit attendance with crude relative risk and risk differences and corresponding 95% confidence intervals.

### Evaluation of the pilot—implementation outcomes

We will also evaluate a comprehensive set of process or implementation outcomes for the pilot period. These outcomes are described in Table 2.

**Table 2.** Implementation outcomes to be evaluated*.

| **Outcome** | **Description** | **Data source** | **Level of analysis** |
| --- | --- | --- | --- |
| Reach | Proportion of eligible clients exposed to toolkit | EMR data; Client IDIs | Facility |
| Fidelity | Correctness with which tools were implemented as intended in each facility | Data collector field and implementation notes, Provider IDIs | Facility |
| Adoption | Uptake, utilization, and intention to try | Provider IDI | Facility and provider |
| Implementation costs based on resource requirements | Staff time by cadre, space, and other resources required to utilize toolkit | Provider IDIs, Authors’ unit cost data | Facility |
| Feasibility | Practical obstacles to use of each tool, if any, suitability for everyday use | Co-creation feedback, Provider IDIs | Facility and provider |
| Appropriateness | Perceived fit, usefulness | Co-creation feedback, Provider IDIs, Client IDIs | Provider and client |
| Acceptability | Satisfaction with toolkit overall and with various components of the toolkit | Co-creation feedback, Provider IDIs, Client IDIs | Provider and client |

\*Adapted from Proctor E, Silmere H, Raghavan R, Hovmand P, Aarons G, Bunger A, Griffey R, Hensley M. Outcomes for implementation research: conceptual distinctions, measurement challenges, and research agenda. *Adm Policy Ment Health*. 2011 Mar;38(2):65-76. doi: 10.1007/s10488-010-0319-7. PMID: 20957426; PMCID: PMC3068522.

### Qualitative analysis methods

For client telephone interviews and provider IDIs, audio recordings will be transcribed verbatim and translated into English. Transcripts will be uploaded to NVivo for qualitative coding and analysis. We will use an inductive-deductive approach, with codebooks designed *a priori* based on the interview guides and the implementation outcomes defined in Table 2, while allowing for new codes to emerge during the coding process. A subset of the transcripts will be coded by two coders and will be assessed for inter-coder reliability using NVivo’s comparison tool. Once agreement is reached, the remaining transcripts will be coded by a single coder. We will conduct a content analysis for each interview type, mapping key themes to implementation outcomes and toolkit component. We will present key emergent themes alongside illustrative quotes. Results will be stratified by district but always presented in aggregate to ensure that no individual will be identifiable.

### Sample size

The anticipated sample size for the primary clinical outcome is based on the expected number of eligible clients who will visit the study and comparison sites during the pilot period and during the six months preceding the pilot. We anticipate that EMR data will be collected for a maximum of 15,000 clients from the pilot sites and a maximum of 15,000 patients from non-pilot comparison sites. Using a two-sided α of 0.05 and assuming a prevalence of missed visits to be 10% at comparison sites, we estimate that this will provide 80% power to detect a relative risk of 1.10.

For the client telephonic interviews, we will enroll up to a maximum of 10 male and 10 female clients per each of the (up to) 8 participating sites (n=160) and will reasonably expect to be able to reach half of these for interviews (40 males, 40 females across all sites). We will enroll up to a maximum of 5 providers of various cadres per participating site for a maximum total sample of 40 provider interviews.

### Dissemination of results

Results of the BRIDGE evaluation will be disseminated in several ways. The primary audience for study findings is the South African Department of Health and its partners, who may use the results to inform programmatic improvements guidelines and procedures during the early treatment period. Broader dissemination will include peer-reviewed publications, presentations at national and international conferences, and policy briefs. Only aggregate and stratified data will be presented to protect confidentiality; no individual participants will be identifiable in any dissemination products.

Depending on results of the evaluation, the BRIDGE tools and/or the full toolkit will also be disseminated as widely as possible. Tools will be posted on relevant websites for free downloading by users, along with orientation materials and guides for utilizing them. We will also work with the Department of Health and with other stakeholders to produce hard copies (posters, handouts) for facilities that wish to use the tools.

### Limitations

We are aware of several limitations that will apply to BRIDGE. First, the very brief pilot implementation period may not be long enough to discern the true impact of the toolkit on the probability of a reduction in treatment interruptions. Second, the sample size will be small, in terms of both the number of study sites and the expected number of clients who will be exposed to the intervention, thus potentially limiting our ability to detect small differences in study outcomes. In this analysis, even small improvements in terms of relative risk may represent large numbers of treatment interruption events prevented at scale. Third, the brief implementation period will only be sufficient to capture providers’ early experiences with the toolkit, not its longer term adoption. This will include the initial period where the providers are still learning and may be using the tools sub-optimally. We will mitigate this to the degree possible by only interviewing providers after implementing at least two weeks. Fourth, providers may exhibit some social desirability bias during interviews, inflating their reported utilization of the tools beyond what was actually done. Client interviews may help to discern this bias should it occur, however.

Finally, as an evaluation of actual use of the toolkit, BRIDGE will not be able to influence or observe intervention fidelity or scale (whether it is used by all providers throughout the pilot period), beyond what providers themselves report to us during interviews. We will thus not be able to distinguish between non-utilization of the tools and non-effectiveness of the tools, if we observe no differences in our primary outcome. This risk of a “false negative” (lack of effect due to poor implementation compliance rather than an ineffective intervention), which is a common shortcoming of implementation research, is in itself a process outcome of the study (“adoption” in Table 2): if providers are unable or choose not to utilize the tools, then they cannot be regarded as successful in practice. The limitations described above will be taken into account as we interpret the results of the pilot study.

### Ethics review

The BRIDGE evaluation protocol was approved by the Institutional Review Board at Boston University Medical Campus (BUMC IRB H-46408, approved March 6, 2026) and the Human Research Ethics Committee (Medical) of the University of the Witwatersrand (251124, approved January 19, 2026). Representatives of the National Department of Health participated in protocol development and will serve as co-investigators for the study. All participants will provide written informed consent prior to qualitative data collection. Medical record review will be conducted under a waiver of informed consent, as approved by the relevant ethical review committees. BRIDGE is registered at Clinicaltrials.gov as NCT07439003.

### Current status of the study

Orientation of facility staff to implement the BRIDGE retention toolkit is taking place in March 2026. The one-month pilot period is anticipated to occur between early April and late May 2026, with start and end dates varying by site.

## Discussion

Despite tremendous improvements in both the medications and service delivery approaches used to treat HIV, treatment interruption and disengagement from care, particularly during the early treatment period, remain prevalent and continue to deter achievement of global and national goals for HIV management. Finding effective, light-touch interventions that can be adopted and scaled within the current resource-constrained environment is thus a very high priority. The BRIDGE retention toolkit incorporates results of multiple prior studies and intervention trials to design and then test a set of five tools to help both clients and providers during the early treatment period. The tools are largely information-based—they aim to provide reminders, guidance, and answers—and are based on South Africa’s existing national guidelines. If some or all of the BRIDGE tools are found to be helpful and/or are associated with a reduction in missed clinic visits, they will comprise a readily scalable and affordable intervention to help address a major barrier in large-scale HIV treatment programs.

## Data availability

Underlying data

No data are associated with this article

Extended data

Extended data are available from https://hdl.handle.net/2144/52738. CC BY-NC 3.0 US Attribution-NonCommercial 3.0 United States.

Supplementary file 1: Study protocol

## Author roles

Sande L: Conceptualization, Methodology, Project Administration, Supervision, Writing—Review and Editing; Maskew M; Conceptualization, Methodology, Project Administration, Supervision, Writing— Review and Editing; Mutanda N: Conceptualization, Methodology, Writing—Review and Editing; Kachingwe E: Conceptualization, Methodology, Writing—Review and Editing; Morgan A: Conceptualization, Methodology, Project Administration, Writing—Review and Editing; Ntjikelane V: Methodology, Project Administration, Writing—Review and Editing; Chiwaye S: Project Administration, Writing—Review and Editing; Benade M: Conceptualization, Methodology, Writing—Review and Editing; Marri AR: Conceptualization, Methodology, Writing—Review and Editing; Malala L: Conceptualization, Supervision, Writing—Review and Editing; Manganye M: Conceptualization, Supervision, Writing— Review and Editing; Rosen S: Conceptualization, Funding Acquisition, Methodology, Project Administration, Writing –Original Draft Preparation, Writing – Review & Editing; Scott N: Conceptualization, Methodology, Project Administration, Writing – Review & Editing.

## Acknowledgments

We would like to acknowledge the guidance and suggestions received from multiple stakeholders during the co-creation process for the BRIDGE Retention Toolkit. These include, in particular, Dr. Anna Grimsrud and Dr. Lynne Wilkinson of the International AIDS Society. We would also like to acknowledge and thank the creators of the WhatsApp Health Coach for allowing us to include it in the BRIDGE toolkit and providing all supporting documentation.

